# Associations between accurate measures of adiposity and fitness, blood proteins, and insulin sensitivity among South Asians and Europeans

**DOI:** 10.1101/2024.09.06.24313199

**Authors:** Pik Fang Kho, Laurel Stell, Shirin Jimenez, Daniela Zanetti, Daniel J Panyard, Kathleen L Watson, Ashish Sarraju, Ming-Li Chen, Lars Lind, John R Petrie, Khin N Chan, Holly Fonda, Kyla Kent, Jonathan N Myers, Latha Palaniappan, Fahim Abbasi, Themistocles L. Assimes

## Abstract

**Objective:** South Asians (SAs) may possess a unique predisposition to insulin resistance (IR). We explored this possibility by investigating the relationship between ‘gold standard’ measures of adiposity, fitness, selected proteomic biomarkers, and insulin sensitivity among a cohort of SAs and Europeans (EURs).

**Methods:** A total of 46 SAs and 41 EURs completed ‘conventional’ (lifestyle questionnaires, standard physical exam) as well as ‘gold standard’ (dual energy X-ray absorptiometry scan, cardiopulmonary exercise test, and insulin suppression test) assessments of adiposity, fitness, and insulin sensitivity. In a subset of 28 SAs and 36 EURs, we also measured the blood-levels of eleven IR-related proteins. We conducted Spearman correlation to identify correlates of steady-state plasma glucose (SSPG) derived from the insulin suppression test, followed by multivariable linear regression analyses of SSPG, adjusting for age, sex and ancestral group.

**Results:** Sixteen of 30 measures significantly associated with SSPG, including one conventional and eight gold standard measures of adiposity, one conventional and one gold standard measure of fitness, and five proteins. Multivariable regressions revealed that gold standard measures and plasma proteins attenuated ancestral group differences in IR, suggesting their potential utility in assessing IR, especially among SAs.

**Conclusion:** Ancestral group differences in IR may be explained by accurate measures of adiposity and fitness, with specific proteins possibly serving as useful surrogates for these measures, particularly for SAs.

## Introduction

Insulin resistance (IR) is characterized by decreased sensitivity to insulin-stimulate glucose uptake in skeletal muscle resulting in a compensatory hyperinsulinemia and several metabolic abnormalities and clinical syndromes, including Type 2 diabetes (T2D) and atherosclerotic cardiovascular disease (ASCVD) (1). On average, individuals of South Asian (SAs) ancestry – generally people whose ancestors originate in India, Sri Lanka, Bangladesh, Nepal, and Pakistan – are more IR, and develop T2D and ASCVD at substantially younger age and lower body mass index (BMI) compared to European ancestry individuals (EURs) (2-4). While some argue that SAs possess a population-specific genetic susceptibility to IR independent of traditional risk factors, the scientific evidence to support this hypothesis is debatable (2-4).

Accurately measuring IR and its primary modifiable determinants – adiposity and cardiorespiratory fitness - in humans is time consuming and expensive (5-7). The Insulin Suppression test (IST) is one of two gold standard methods for direct measurement of insulin-stimulated glucose uptake that yields a measure of IR with unequivocal physiological interpretation but requires intensive monitoring over ∼3-4 hours to complete (5). Adiposity is commonly estimated by documenting body mass index (BMI) and waist circumference (WC) but these measures do not accurately reflect the volume and distribution of body fat in relation to other types of tissues (8). Physical fitness is usually assessed through self-report questionnaires of physical activity which are susceptible to recall and information bias (9). Gold standard measures such as dual energy x-ray absorptiometry (DXA) and maximal VO_2_ (VO_2_ max) may provide a more accurate assessment of adiposity and physical fitness, respectively, and may better explain differences in IR between populations at variable risk (8, 10-12). Moreover, recent advancements in the field of plasma proteomics have enabled the identification of proteomics biomarkers that can predict gold standard measure of IR more accurately than the clinical variables (13). The role of such proteins in explaining differences in insulin sensitivity between populations is unclear.

We hypothesized that differences in IR between SAs and EURs may be at least partially attributed to the usage of suboptimal assessments of adiposity, fitness, and IR. To test this hypothesis, we measured these health traits using both ‘conventional’ and, more accurate, ‘gold standard’ approaches in a modest number of healthy SAs and EURs and assessed whether the gold standard measures might better explain ancestral differences in insulin sensitivity between SAs and EURs. In addition, we assessed whether the levels of eleven proteins in plasma previously robustly associated with IR might explain differences in insulin sensitivity between the two groups (13).

## Materials and Methods

### Study Population and Design

We recruited generally healthy individuals between 19 and 75 years of age who were living in the San Francisco Bay Area and reported that all four of their biological grandparents originated either from South Asian countries (Afghanistan, Bangladesh, Bhutan, India, Maldives, Pakistan, Nepal and Sri Lanka) or European countries. Volunteers responded to local (on campus) and regional (off campus) in-print and email advertisements about the study, and eligible participants were enrolled between 2012 and 2015. We excluded individuals with any type of diabetes, complications of ASCVD, active cancer, any terminal disease, renal dysfunction, liver dysfunction, anemia, systolic blood pressure >160 mm Hg, heart block, or bradyarrhythmias. We also excluded pregnant or lactating women and individuals receiving treatment with glucose-lowering medications.

Participants underwent a 2-day testing protocol. On Day 1 of the protocol, volunteers completed informed consent and then underwent a DXA scan to document regional body fat followed by an exercise treadmill test to document VO_2_ max. They also completed a questionnaire on their lifetime physical activity after the age of 25 years. On Day 2 of the protocol, participants underwent an insulin suppression test (IST) and filled out additional questionnaires documenting their medical history, current prescribed and over-the-counter medications, and their aerobic physical activity in the seven days before the IST.

Additionally, a standardized protocol was used to process and bank fasting blood for future biomarker analysis. The protocol was reviewed and approved by the Institutional Review Board at the Stanford University School of Medicine and all participants provided informed consent.

### Clinical Measures related to Adiposity

Adiposity was measured using conventional and gold standard measures. Conventional measures included weight, height, and waist circumference (WC). On the day of the IST, weight was measured while participants were wearing light clothing and no shoes. Height was measured by a stadiometer. Body mass index (BMI) was calculated by dividing weight in kilograms by the square of height in meters. WC was measured at the midpoint between the lower rib cage and the upper iliac crest in mid respiration while participants were standing. The gold standard measures of adiposity were derived through DXA. We used a Lunar iDXA scanner (GE Healthcare) to perform regional body composition analysis of all volunteers that included quantification of fat mass, fat-free mass, and bone mineral content. With this scanner, android and gynoid regions-of-interest (ROIs) are automatically placed and ratios are automatically calculated. The android ROI includes the area between the pelvis cut line extending upwards to include 20% of the distance between the pelvis and neck cut lines. The arm cut lines when in the normal position for a total body scan define lateral boundaries. The gynoid ROI is positioned with the upper boundary positioned below the pelvis cut line a distance equal to 1.5 times the height of the android region. The lower boundary is located a distance 3.5 times the height of the android region from the pelvis cut line. Lateral boundaries are the outer leg cut lines.

### Clinical Measures related to Physical Activity and Fitness

Physical activity was measured using conventional and gold standard measures. Conventional measures included self-report questionnaires. We assessed lifetime physical activity using the Lifetime Physical Activity Questionnaire (14) and physical activity within the last week using the short form of the International Physical Activity Questionnaire (IPAQ) (15). The IPAQ data were scored using the continuous score protocol in which Metabolic Equivalent of Task (MET)-minutes for each study participant’s activity were calculated as per standardized guidelines. The gold standard measure of fitness consisted of a cardiopulmonary exercise treadmill test (CPET) where we implemented a standardized symptom-limited, individualized ramp treadmill testing protocol combined with a metabolic cart to determine maximal oxygen consumption (i.e., VO_2_ max) (16). Briefly, participants filled out a short one-page questionnaire about physical activities on the day of the CPET designed to predict a participants peak MET level. This estimate was then used to tailor the “ramp” protocol in which small increments of work rate occurred at intervals of <10 to 60 seconds to yield a fatigue-limited exercise duration of ∼8 to 12 minutes. Effort was assessed using the Borg Rating of Perceived Exertion and the test was considered successful when a scale of 17 or more was reported and the participant indicated that the test be stopped due to peak exercise leg weakness or intolerable dyspnea on exertion. Importantly, all except one participant for this study had fasting plasma glucose levels within the normal range (< 125 mg/dL), with no history of diabetes, either treated or untreated. These criteria ensured that the VO_2_ max measurements reflected true cardiorespiratory fitness, free from potential confounding effects of hyperglycemia.

### Insulin Suppression Test

We performed an IST to directly measure insulin-stimulated glucose disposal (17). Briefly, intravenous (IV) access was obtained in each arm of the participants after an overnight fast. One IV was used for administering a continuous infusion of octreotide acetate (0.27 μU/m^2^/min), insulin (32 mU/m^2^/min), and glucose (267 mg/m^2^/min) at a constant rate for up to three hours, while the other IV was used for collecting blood samples. Blood samples were drawn every 30 minutes (min) until 150 min for plasma glucose and then every 10 min until 180 min for plasma insulin and glucose. We averaged the four insulin and glucose values obtained from 150 to 180 min to calculate the steady-state plasma insulin (SSPI) and steady-state plasma glucose (SSPG) concentrations. SSPI concentrations were comparable among participants with identical glucose infusion rates making the magnitude of the SSPG an accurate estimate of one’s insulin-mediated glucose disposal, with a higher SSPG indicating a more insulin-resistant individual.

### Plasma proteomic measures

We employed the proximity extension assay developed by OLINK, utilizing the first version of EXPLORE platform to quantify up to 1,471 proteins across four 384-plex panels (Cardiometabolic, Inflammation, Neurology and Oncology) in plasma samples from a subset of SA and EUR individuals. This analysis is part of a broader study encompassing 1,012 participants from various research protocols at Stanford, including assessments of insulin sensitivity using the IST (to be reported separately). Samples from 18 SAs and 5 EURs were inadvertently lost during a reorganization of freezers, leaving 28 SAs and 36 EURs in this analysis. Our analysis focused on 11 plasma proteins which were previously shown to predict gold standard measures of insulin sensitivity in two European cohorts (RISC and ULSAM). These proteins included: Fatty acid binding protein 2 (FABP2), Fatty acid binding protein 4 (FABP4), Insulin-like growth factor binding protein 1 (IGFBP1), Insulin-like growth factor binding protein 2 (IGFBP2), Inhibin beta C chain (INHBC), Leptin (LEP), Reticulon 4 receptor (RTN4R), Secretoglobin family 3A member 2 (SCGB3A2), Adhesion G protein-coupled receptor G1 (ADRG1), Integrin subunit alpha V (ITGAV), and Lipoprotein lipase (LPL). The Unitprot ID for each protein can be found in **Supplementary Table 1**.

The protein profiling was performed by the Psomagen service lab following OLINK’s standard operating procedure. Quality control for the EXPLORE proteomic data was applied to all samples profiled at Stanford, which included samples from 28 SAs and 36 EURs in this study, using the “OlinkAnalyze” package in R, which is developed and maintained by the Olink Proteomics Data Science Team. First, we identified outliers within each protein panel through (1) principal component analysis and (2) assessment of median and inter-quartile range (IQR) values for Normalized Protein eXpression (NPX) across all samples measured. Data points were excluded if they (1) exhibited a standardized PC1 or PC2 value more than five standard deviations from the mean (in standardized PCA, this mean is zero), or (2) had a median NPX or IQR of NPX more than five standard deviations from their respective mean values. Further data points with QC or assay warnings were also removed.

### Statistical Analysis

We first imputed missing variables using predictive mean matching (pmm) as implemented in the R package ‘mice’(18) to minimize the loss of observations and maximize the power of our analyses. While a small number of missing protein levels were imputed among subjects with protein levels, we did not attempt to impute all protein levels in subjects who did not have any proteins measured.

Descriptive statistics are presented as mean +/-standard deviation for continuous variables and as count and percentages for categorical variables. Differences in baseline characteristics between EURs and SAs were compared using a t-test for continuous variables and a chi-square test for categorical variables. We computed Spearman correlations and P values between each measure and SSPG to identify variables significantly correlated with SSPG, adopting Bonferroni adjusted significance threshold 0.05/30=0.00167 to account for the testing of 30 measures. This analysis included all 87 participants for non-protein variables and a subset of 64 participants for protein variables. The Spearman correlation was chosen due to its robustness to outliers and suitability for non-normally distributed data. Variables with significant correlations at the Bonferroni-adjusted threshold were identified as candidate predictors for further analysis regarding their role in SSPG differences between EUR and SA participants.

SSPG values were log-transformed to approximate a normal distribution prior to linear regression analysis of SSPG (**Supplementary Figure 1**). A base model with covariates age, sex and ancestral group was used for comparison. The first goal was to study whether adding selected groups of variables to this model attenuated the significance of the ancestral group variable. We considered three groups of variables from among those significantly correlated with SSPG: a) conventional measures of adiposity and physical activity, b) gold standard measures of adiposity and cardiorespiratory fitness, c) plasma proteins. The second goal was to estimate the extent to which different models accounted for the variability in SSPG. We performed multivariable linear regressions for the models described above, excluding the ancestral group variable, to evaluate the variance explained (R^2^) and residual standard error (SE) of SSPG in the combined ancestral groups and separately for each ancestral group. Lastly, to investigate the mediation path of select proteins significantly correlated with SSPG, we computed their Pearson correlations with the gold standard measures of adiposity and cardiorespiratory fitness. All analyses were performed using R.

## Results

We enrolled 107 participants into the study but only 87 completed the testing protocol, including 46 SAs and 41 EURs. Among those completing the protocol, 43% completed it within one month, 76% within three months, 87% within six months, and 99% within a year. **Table 1** summarizes the characteristics of the study participants, stratified by ancestral group. SAs were, on average, younger, had higher percentages of total adipose tissue, as well as adipose tissue within their trunk and android regions. SAs also had a lower lifetime recreational physical activity and reported lower amounts of physical activity in the week before their IST. Significant differences between ancestral groups in plasma proteins related to insulin sensitivity were also observed, with EURs showing higher levels of IGFBP1, IGFBP2, LPL, SCGB3A2 and ITGAV, but lower levels of LEP and INHBC.

**Table 1.** Characteristics of 41 European and 46 South Asian study participants.

|  |  |  |  |  |  |
| --- | --- | --- | --- | --- | --- |
|  | **Europeans** |  | **South Asians** |  |  |
| **Variables** | **Mean** | **SD** | **Mean** | **SD** | **P value** |
| **Demographics** |  |  |  |  |  |
| Age | 52.1 | 12.2 | 43.0 | 8.3 | <0.001 |
| Age* | 53.7 | 12.1 | 42.8 | 8.4 | <0.001 |
| Sex (N, %) |  |  |  |  | 0.36 |
| Female | 21 | 51.2 | 18 | 39.1 |  |
| Male | 20 | 48.8 | 28 | 60.9 |  |
| Sex (N, %)* |  |  |  |  | 0.592 |
| Female | 19 | 52.8 | 12 | 42.9 |  |
| Male | 17 | 47.2 | 16 | 57.1 |  |
| **Adiposity** |  |  |  |  |  |
| *Conventional measures* |  |  |  |  |  |
| BMI | 24.8 | 3.9 | 25.5 | 3.3 | 0.355 |
| Waist circumference | 88.4 | 11.6 | 88.5 | 12.7 | 0.995 |
| *DXA-derived 'gold standard' measures* |  |  |  |  |  |
| % of total body fat | 29.6 | 9.7 | 33.4 | 6.5 | 0.032 |
| % of total body fat in trunk region | 51.1 | 7.6 | 53.9 | 6.9 | 0.069 |
| % of total body fat in leg region | 34.3 | 6.7 | 32.3 | 6.2 | 0.145 |
| % of total body fat in arm region | 10.0 | 1.4 | 9.7 | 1.4 | 0.434 |
| % of total body fat in gynoid region | 19.5 | 3.5 | 18.4 | 3.0 | 0.104 |
| % of total body fat in android region | 8.8 | 2.1 | 9.6 | 1.7 | 0.058 |
| % trunk (region) that is fat | 30.9 | 10.8 | 37.1 | 7.4 | 0.002 |
| % leg (region) that is fat | 29.9 | 10.9 | 31.5 | 7.6 | 0.429 |
| % arm (region) that is fat | 27.9 | 11.4 | 29.8 | 9.1 | 0.389 |
| % android (region) that is fat | 35.4 | 13.1 | 43.6 | 8.4 | 0.001 |
| % gynoid (region) that is fat | 36.4 | 12.5 | 39.3 | 8.7 | 0.214 |
| Android to gynoid fat ratio | 0.5 | 0.2 | 0.6 | 0.2 | 0.073 |
| **Physical Activity and Fitness** |  |  |  |  |  |
| *Conventional measures* |  |  |  |  |  |
| LY Rec kcal/week | 2643 | 1896 | 2017 | 1365 | 0.078 |
| LT Rec kcal/week | 2000 | 1368 | 1286 | 884 | 0.004 |
| LT Occ kcal/week | 7946 | 2249 | 7386 | 2096 | 0.232 |
| PA MET score last week | 2445 | 1658 | 1376 | 800 | <0.001 |
| *Cardiopulmonary Exercise Treadmill Test derived 'gold standard' measure* |  |  |  |  |  |
| VO2 max | 36.92 | 10.3 | 34.78 | 6.19 | 0.237 |
| **Protein measures in plasma\*** |  |  |  |  |  |
| FABP2 | 0.32 | 0.88 | 0.06 | 0.82 | 0.227 |
| FABP4 | -0.66 | 0.73 | -0.44 | 0.61 | 0.22 |
| IGFBP1 | 1.09 | 1.1 | 0.17 | 1.05 | 0.001 |
| IGFBP2 | 0.66 | 0.59 | -0.06 | 0.64 | <0.001 |
| LEP | -1.59 | 1.94 | -0.41 | 1.23 | 0.007 |
| LPL | 0.22 | 0.61 | -0.19 | 0.53 | 0.006 |
| SCGB3A2 | 0.63 | 0.77 | 0.22 | 0.76 | 0.037 |
| ADGRG1 | 0.11 | 0.87 | -0.23 | 0.49 | 0.075 |
| ITGAV | 0.06 | 0.2 | -0.16 | 0.21 | <0.001 |
| RTN4R | -0.22 | 0.38 | -0.09 | 0.46 | 0.246 |
| INHBC | -0.44 | 0.66 | -0.04 | 0.43 | 0.008 |
| **Insuling Sensitivity** |  |  |  |  |  |
| Fasting plasma glucose | 96.3 | 9.2 | 97.9 | 8.2 | 0.388 |
| SSPG | 94.6 | 46.9 | 130.2 | 57.7 | 0.002 |
| SSPG* | 91.0 | 43.5 | 132.8 | 62.7 | 0.003 |
| Data are mean ± standard deviation (SD) (range) unless otherwise noted. *Subset with plasma proteins: 36 Europeans, 28 South Asians. SSPG: steady state plasma glucose derived from the insulin suppression test. concentration is a direct measure of insulin resistance (IR) where a higher SSPG concentration indicates greater IR compared to a lower SSPG concentration. BMI: body mass index; LY Rec kcal/week: Last-year recreational physical activity kcal/week; LT Rec kcal/week: Lifetime recreational physical activity kcal/week; LT Occ kcal/week: Lifetime occupational physical activity kcal/week; PA MET score last week: Physical activity metabolic task equivalent score last week; SSPG, steady-state plasma glucose; and VO2 max, amount of oxygen utilized during a cardiopulmonary exercise test with maximal effort; ADGRG1: Adhesion G Protein-Coupled Receptor G1; FABP2: Fatty Acid Binding Protein 2; FABP4: Fatty Acid Binding Protein 4; IGFBP1: Insulin Like Growth Factor Binding Protein 1; IGFBP2: Insulin Like Growth Factor Binding Protein 2; INHBC:Inhibin Subunit Beta C; ITGAV: Integrin Subunit Alpha V; LEP: Leptin; LPL: Lipoprotein Lipase; RTN4R: Reticulon 4 Receptor; SCGB3A2: Secretoglobulin Family 3A Member 2 | | | | | |

The Spearman correlation analysis identified 16 variables significantly correlated with SSPG at Bonferroni correction (**Figure 1**). The variables that exhibited either a significant positive or negative correlation with SSPG included one conventional (BMI) and eight DXA derived measures of adiposity, one conventional (PA MET score last week from IPAQ) and one gold standard (VO_2_ max) of physical fitness and five proteins (**Supplementary Figure 2**). Among the conventional measures, BMI had the strongest correlation, which was only the tenth strongest overall. For subsequent multivariable regression analyses, we retained only percent trunk that is fat variable among eight adiposity variables having significant Spearman correlation with SSPG given this variable had the strongest correlation with SSPG and was moderately to strongly correlated with other adiposity measures (r=0.45–0.99). Among the proteins correlated with SSPG, we excluded from the linear regression FABP4, which was strongly correlated with LEP (r=0.75), and IGFBP1, which was strongly correlated with IGFBP2 (r=0.73) and retained INHBC, LEP, IGFBP2.

**Figure 1.**
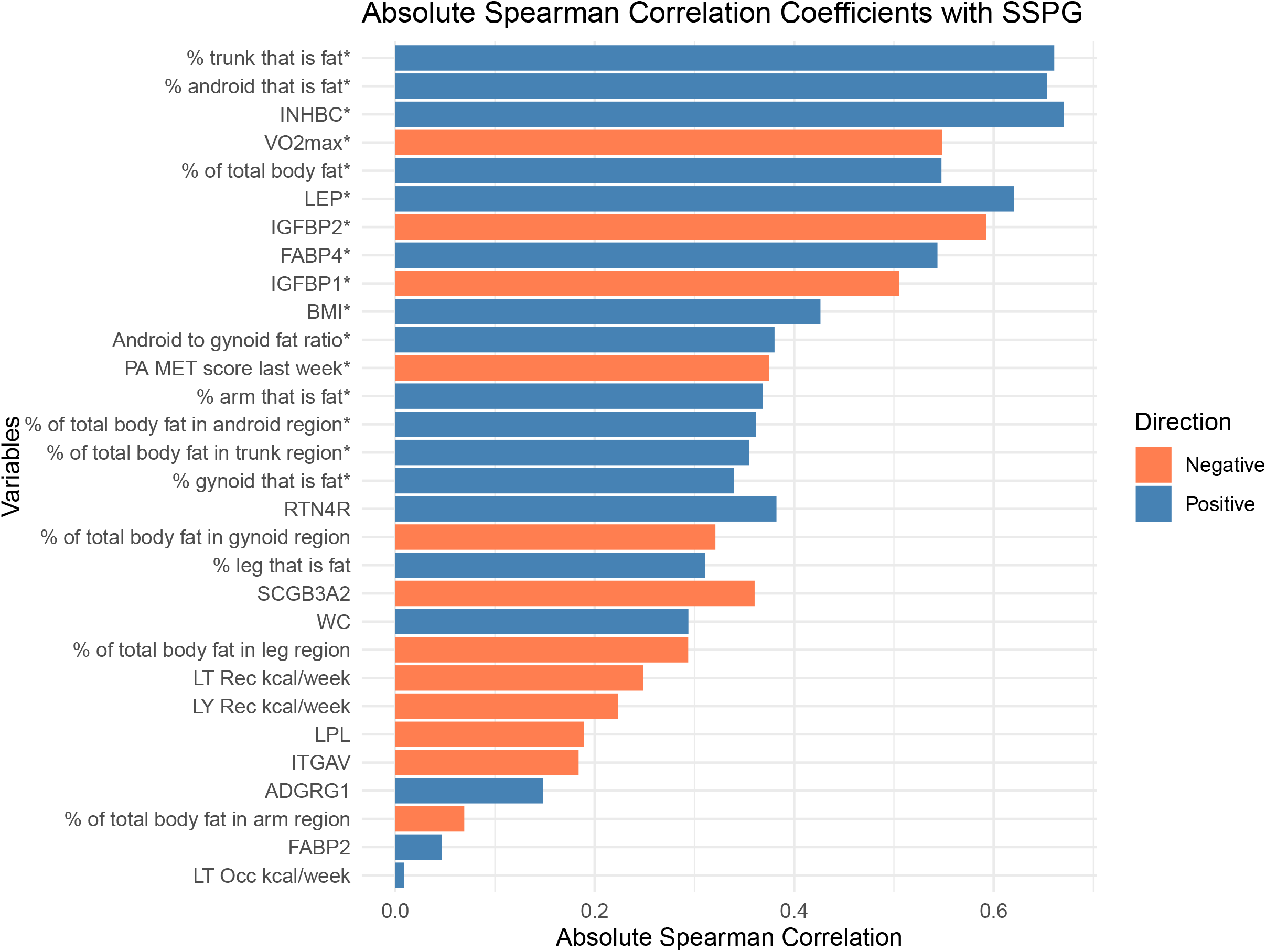
Absolute Spearman correlation with SSPG, sorted by P values. The color represents the direction of the correlations, with blue indicating positive correlations and orange indicating negative correlations. The length of each bar represents the absolute value of the Spearman correlation coefficients. Variables significantly correlated with SSPG at Bonferroni-adjusted significance levels (P < 0.00167) are marked with an asterisk (*).

To make comparisons between models more justifiable, all were fit using the subset of participants with protein measures. In linear regression of log-transformed SSPG with covariates age, sex and ancestry, difference in mean SSPG levels between ancestry groups was strongly significant (ß=0.471, SE=0.125, P=0.0004). Adding conventional measures, BMI and PA MET score to this model reduced the ß coefficient for ancestral group by 39% although its significance persisted (ß=0.286, SE=0.119, P=0.019). The difference between ancestral groups was further reduced and was no longer significant when gold standard measures (percent trunk that is fat, VO_2_ max) (ß=0.142, SE=0.116, P=0.228) or the three retained IR-related proteins levels in plasma (ß=0.069, SE=0.114, P=0.012) were incorporated (**Figure 2; Supplementary Table 2**).

**Figure 2.**
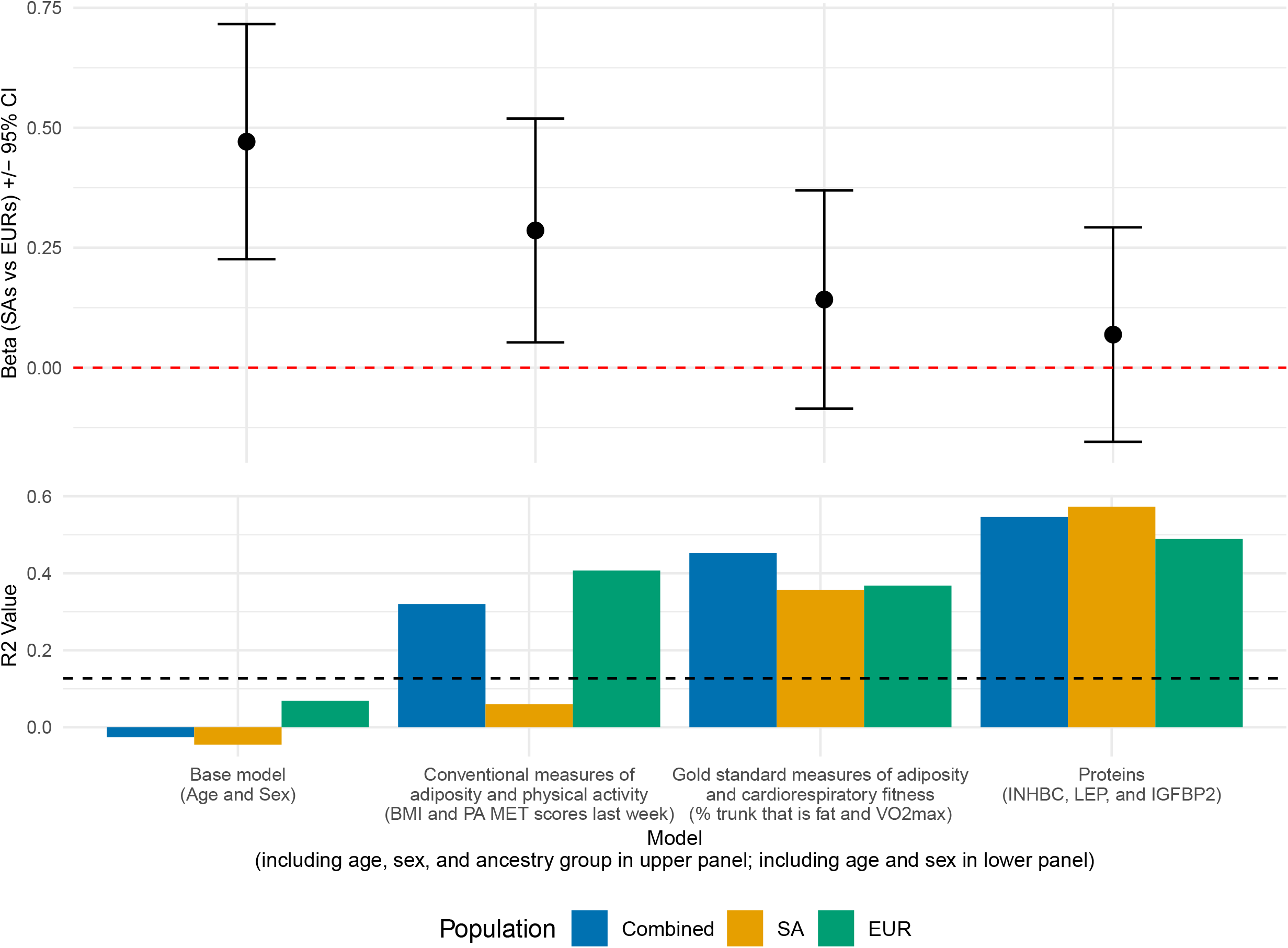
Results from multivariable regression of log(SSPG) with selected sets of covariates, fitting only participants with protein measures. The top panel displays the beta coefficients (+/-95% confidence interval) for the ancestral group variable (SA vs EUR) in each model: base model (age and sex), conventional measures of adiposity and physical activity (BMI and PA MET scores last week), gold standard measures of adiposity and fitness (percent trunk that is fat and VO_2_ max), and proteins (INHBC, LEP, IGFBP2). The red dashed line indicates a beta coefficient of zero. The bottom panel displays the adjusted R^2^ values of the same models, excluding ancestral group variable, for the combined ancestral group (EURs and SAs) in blue, as well as separately for SA (orange) and EUR (green) participants. The black dashed line at 0.127 indicates the adjusted R^2^ from the base model with age, sex, and ancestral group.

To assess R^2^ for SSPG by different models, we repeated multivariable linear regressions described above without the ancestral group variable. This analysis evaluated the R^2^ for SSPG in the combined as well as in the SAs and EURs separately. The R^2^ values when using conventional measures of adiposity and physical activity (SA: 0.06; EUR: 0.41; combined ancestral groups: 0.32) show that the combination of these measures was much more strongly associated with SSPG in EUR participants compared to SA participants. The R^2^ values when using gold standard measures of adiposity and cardiorespiratory fitness (SA: 0.36; EUR: 0.37; combined ancestral groups: 0.45) was about the same in both ancestry groups—and also comparable to the value in EUR for the conventional measures. The protein model showed the highest R^2^ values (SA: 0.57; EUR: 0.49; combined ancestral groups: 0.55) and might be more strongly associated with SSPG in SA than in EUR. The residual SEs of these fits showed similar trends (**Supplementary Table 2**). Additionally, we observed significant correlations between gold standard measures of adiposity and fitness and the five proteins that were significantly correlated with SSPG (**Table 2**). Notably, percent trunk that is fat showed strong positive correlation with FABP4 and LEP (r>0.7). Concurrently, these two proteins showed very strong negative correlations with VO_2_ max. IGFBP1 and IGFBP2 were significantly correlated with percent trunk that is fat but not with VO_2_ max.

**Table 2.** Pearson correlations between gold standard measures and plasma protein, ordered by P value within each gold standard measure.

|  |  |  |  |
| --- | --- | --- | --- |
| **Gold standard measures** | **Proteins** | **Pearson correlation** | **P_value** |
| % trunk (region) that is fat | LEP | 0.853 | <1E-15 |
| % trunk (region) that is fat | FABP4 | 0.775 | 5.84E-14 |
| % trunk (region) that is fat | INHBC | 0.657 | 3.61E-09 |
| % trunk (region) that is fat | IGFBP1 | -0.461 | 1.25E-04 |
| % trunk (region) that is fat | IGFBP2 | -0.435 | 3.30E-04 |
| VO<sub>2</sub> max | LEP | -0.814 | <1E-15 |
| VO<sub>2</sub> max | FABP4 | -0.729 | 8.70E-12 |
| VO<sub>2</sub> max | INHBC | -0.451 | 1.85E-04 |
| VO<sub>2</sub> max | IGFBP2 | 0.129 | 3.10E-01 |
| VO<sub>2</sub> max | IGFBP1 | 0.129 | 3.11E-01 |
| VO<sub>2</sub> max, amount of oxygen utilized during a cardiopulmonary exercise test with maximal effort; FABP4: Fatty Acid Binding Protein 4; IGFBP1: Insulin Like Growth Factor Binding Protein 1; IGFBP2: Insulin Like Growth Factor Binding Protein 2; INHBC: Inhibin Subunit Beta C; LEP: Leptin. |  |  |  |

## Discussion

In this study, we evaluated and compared the relationship between both conventional and gold standard measures of adiposity and fitness, as well as specific plasma proteins, with SSPG. We found the strongest correlates of SSPG to be multiple DXA-derived measures, VO_2_ max, and multiple plasma proteins previously robustly linked to IR. Correlations with SSPG for these more accurate measures were notably higher when compared to conventional measures such as BMI and questionnaire-based measures of physical activity. Including either the gold standard measures of adiposity and fitness or the plasma proteins in multivariate linear regressions of SSPG attenuated differences in SSPG between ancestral groups and made these difference non-significant, suggesting that disparities in IR between SAs and EURs might be at least partially due to the availability and accuracy of these measures.

The observed differences in the strength of associations and explanation of variance of various regression models between EUR and SA participants suggest that tailored approaches may be necessary for effective management and intervention in diverse populations. In our study, gold standard measures of adiposity and fitness, such as percent trunk that is fat and VO_2_ max, explained more of the variance in SSPG in EUR participants than in SAs. Conversely, five plasma proteins explained more of the variance in SSPG in SA participants. These findings underscore the importance of considering adiposity, fitness and protein biomarkers when assessing insulin sensitivity in different ancestral groups. If validated, this understanding suggests the need for population-specific strategies to effectively address insulin resistance, considering the unique physiological and metabolic profiles of different ancestral groups.

Prior studies have reported that SAs have a greater degree of IR and face a higher risk of IR-related complications compared with EURs (2, 19, 20). However, few have documented differences between SAs and EURs using direct ‘gold standard’ measures of insulin sensitivity. Given the impractical nature of conducting a direct measure of insulin sensitivity in large numbers, most large-scale epidemiologic studies have generally made use of more easily obtainable surrogate measures of IR (1, 21). Not surprisingly, the four studies that have published such data have consisted of cross-sectional analyses of small cohorts (n =10 to 29 South Asians) which have nevertheless demonstrated a significantly lower degree of insulin sensitivity in SAs compared to whites independent of age, sex, BMI, and physical activity (22-25). Of these, three assessed fat mass using “doubly indirect” methods such as waist circumference, BMI, skin folds, and bioelectrical impedance, while the fourth used the more reliable “singly indirect” method of densitometry (22-25). Although this latter study suggested persistent lower insulin sensitivity in SAs compared to whites at various percentages of fat, a test of the statistical significance of these differences was not reported (25). None of these studies measured and adjusted for VO_2_ max although differences in VO_2_ max and caliper-based measure of adiposity explained differences in a surrogate measure of IR, Homeostatic Model Assessment for Insulin Resistance (HOMA-IR), in a fifth study of interest (26). To our knowledge, our study is the first to use gold-standard measures for IR, adiposity, and cardiorespiratory fitness to assess differences in IR.

The increased IR among SAs compared to EURs may be driven by multiple related ancestral group differences involving the handling of the transformation of excess calories into adipose tissue. First, the overall degree of adiposity may differ given prior studies have documented higher percent body fat at each level of BMI among SAs compared to EURs (27, 28). Second, SAs may be more prone than EURs to store surplus energy within the visceral adipose tissue (VAT) compartment, rather than subcutaneous adipose tissue (SAT), increasing risk through the negative metabolic effects of ectopic fat on the liver and other intra-abdominal organs (27, 29-31). A newly emerging but related hypothesis suggests that the metabolic outcomes of obesity might also be linked to a reduced ability to store energy in the SAT compartment (32, 33). This hypothesis is supported by the identification of a heritable ‘favorable adiposity phenotype’ through recent genome-wide association studies and the possibility that the frequency of such favorable genetic variants is decreased among SAs, although this conjecture still requires further, larger genetic studies for substantiation (32, 33). Furthermore, it is important to highlight that the evidence suggesting stronger correlations between VAT and IR, as compared to SAT and IR, using both proxy and direct measures, remains inconclusive (34, 35). Lastly, a recent study involving two UK-based cohorts that underwent abdominal magnetic resonance imaging (MRIs) failed to confirm an elevated VAT to SAT ratio and intrahepatic fat percentage among SAs compared to EURs (36).

Lower levels of physical activity among SAs compared to EURs may also be contributing to differences in insulin sensitivity (37-40). Physical activity has both immediate and long-term effects on insulin sensitivity that may be mediated through reductions in VAT and not be fully captured through a measure of physical fitness such as V02 max which largely reflects a combination of both peripheral adaptations within skeletal muscle as well as adaptations centrally involving more mechanical aspects of the cardiovascular system such as cardiac output. We attempted to capture both long-term and short-term effects on insulin sensitivity through the CPET and our self-reported physical activity questionnaires but found that the CPET-derived VO2 max was the most strongly correlated measure to SSPG with no incremental benefit provided by our physical activity questionnaire data.

We found plasma levels of INHBC, FABP4, and LEP to be moderately to highly correlated with both percent trunk that is fat and VO_2_ max, suggesting that these proteins are integrally linked with both adiposity and fitness. In contrast, IGFBP2 and IGFBP1 were only significantly correlated with percent trunk that is fat, suggesting their relationship with SSPG is primarily mediated through adiposity rather than fitness. The role of these five candidate proteins in metabolic regulation is reasonably well established. INHBC, a member of the transforming growth factor-beta family, regulates the secretion of follicle-stimulating hormone, hypothalamic, pituitary, gonadal hormones, and insulin (41). Additionally, INHBC is highly expressed in the liver (https://gtexportal.org/home/gene/INHBC). LEP, primarily secreted by adipose tissue, is a key regulator of energy balance and glucose metabolism (42). IGFBP2 is known to regulate insulin and glucose metabolism and has been found to be significantly increased following bariatric surgery in parallel to the improvement in insulin sensitivity (43, 44). Similarly, IGFBP1 also regulates insulin and glucose metabolism with higher levels often observed in individuals with higher insulin sensitivity (45). Lastly, FABP4 is primarily found in macrophages and adipose tissue, where it plays a crucial role in controlling the storage and breakdown of fatty acids and serves as a key mediator of inflammation (42). Given the moderate-to-high correlation between these proteins with gold standard measures of adiposity and/or fitness, they may serve as proxies for gold standard measures of adiposity and fitness acquired through DXA scan and CPET. If further replicated, the routine measurement of these proteins has the potential to substantially improve the identification of individuals at risk with a fraction of the cost of more costly and less accessible gold standard procedures.

Our study has certain limitations worthy of mention. First, the study population consisted of relatively healthy volunteers from the San Francisco Bay Area, which may limit the generalizability of our findings. The observed ordering of measures and models by strength of correlation, differences between ancestry groups and explanation of variability in SSPG might not persist in a different sample. The SA sample also included a large geographic area covering various countries, and there may be heterogeneity within the South Asian sub-groups that is not captured. While we reported that plasma protein and gold standard measures of adiposity and cardiorespiratory fitness appear to correlate with differences in insulin sensitivity between EURs and SAs, we also acknowledge that these results are based on the specific characteristics and potential recruitment biases of the study sample. Although the availability of gold standard measures overall is a key strength, the cost and time required to perform these studies is great, which restricted the sample size and power of our study. Among the strongest correlates for SSPG, multiple DXA-derived measures of adiposity were highly correlated with each other. We retained only the percent trunk that is fat in the study due to its strongest association with SSPG, but we cannot discard the possibility that other adiposity measures may ultimately serve as better correlates to SSPG in the general population. Similarly, we cannot determine whether plasma protein LEP or FAB4 is more explanatory, or IGFBP1 versus IGFBP2. Furthermore, the protein measures are relative quantifications, and our findings require replication and validation in larger sample sizes with assays that can deliver absolute levels before they can be integrated into clinical care. Lastly, the cross-sectional design of the study precludes causal inferences. Future studies with larger sample sizes and longitudinal designs are warranted to validate our findings and to explore the causal pathways linking these candidate predictors to SSPG within and across ancestral groups.

In conclusion, our study demonstrated that accurate measures of adiposity and cardiorespiratory fitness, as well as plasma proteins, may help to explain differences in SSPG between EUR and SA participants as well as variability within these ancestry groups. These measures may provide a more comprehensive understanding of insulin sensitivity differences between EUR and SA participants, with substantially more explanatory power than BMI. Our findings underscore the importance of considering multiple biological factors in the assessment of insulin resistance and support the potential utility of specific plasma proteins as biomarkers for personalized intervention strategies.

## Data Availability

All data produced in the present work are contained in the manuscript

## Funding and Assistance

This study was supported by grants from the National Institutes of Health K23DK088942 and R01DK11418.

## Author Contributions and Guarantor Statement

K.N.C., H.F., K.K., J.N.M., L.P., F.A., and T.L.A. were involved in the conception, design, and conduct of the study; P.K., L.S., S.J., F.A., and T.L.A were involved in the analysis and interpretation of the results. P.K., L.S., A.S., F.A., and T.L.A. wrote the initial drafts of the manuscript, and all authors edited, reviewed, and approved the final version of the manuscript. T.L.A. is the guarantor of this work and, as such, had full access to all the data in the study and takes responsibility for the integrity of the data and the accuracy of the data analysis.

