## Supplementary Materials for "Associations between accurate measures of adiposity and fitness, blood proteins, and insulin sensitivity among South Asians and Europeans"

### Figures

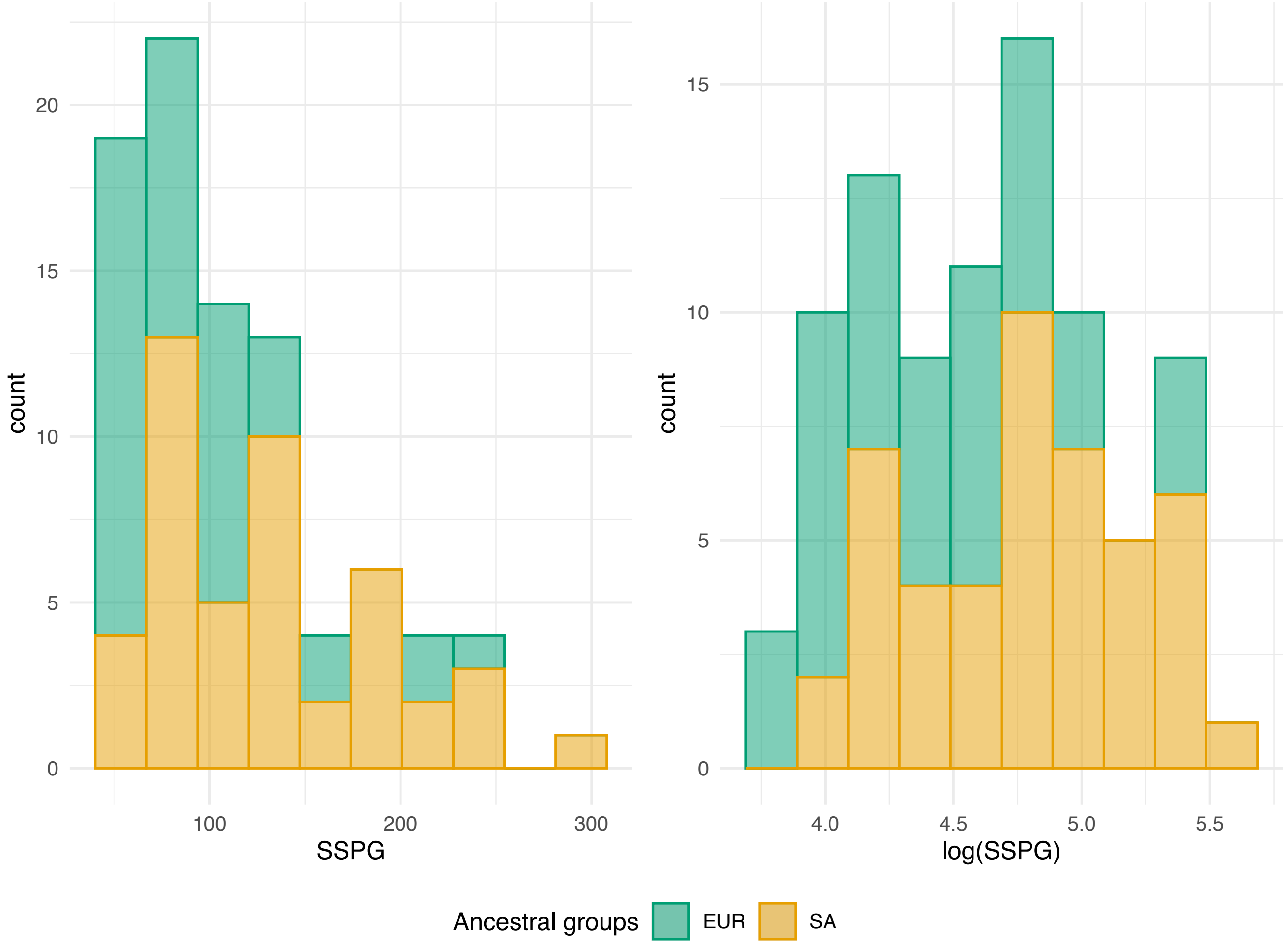


**Supplementary Figure 1. Distribution of SSPG and log(SSPG) values.** The colors indicate the different ancestral groups: EUR (green) and SA (orange).

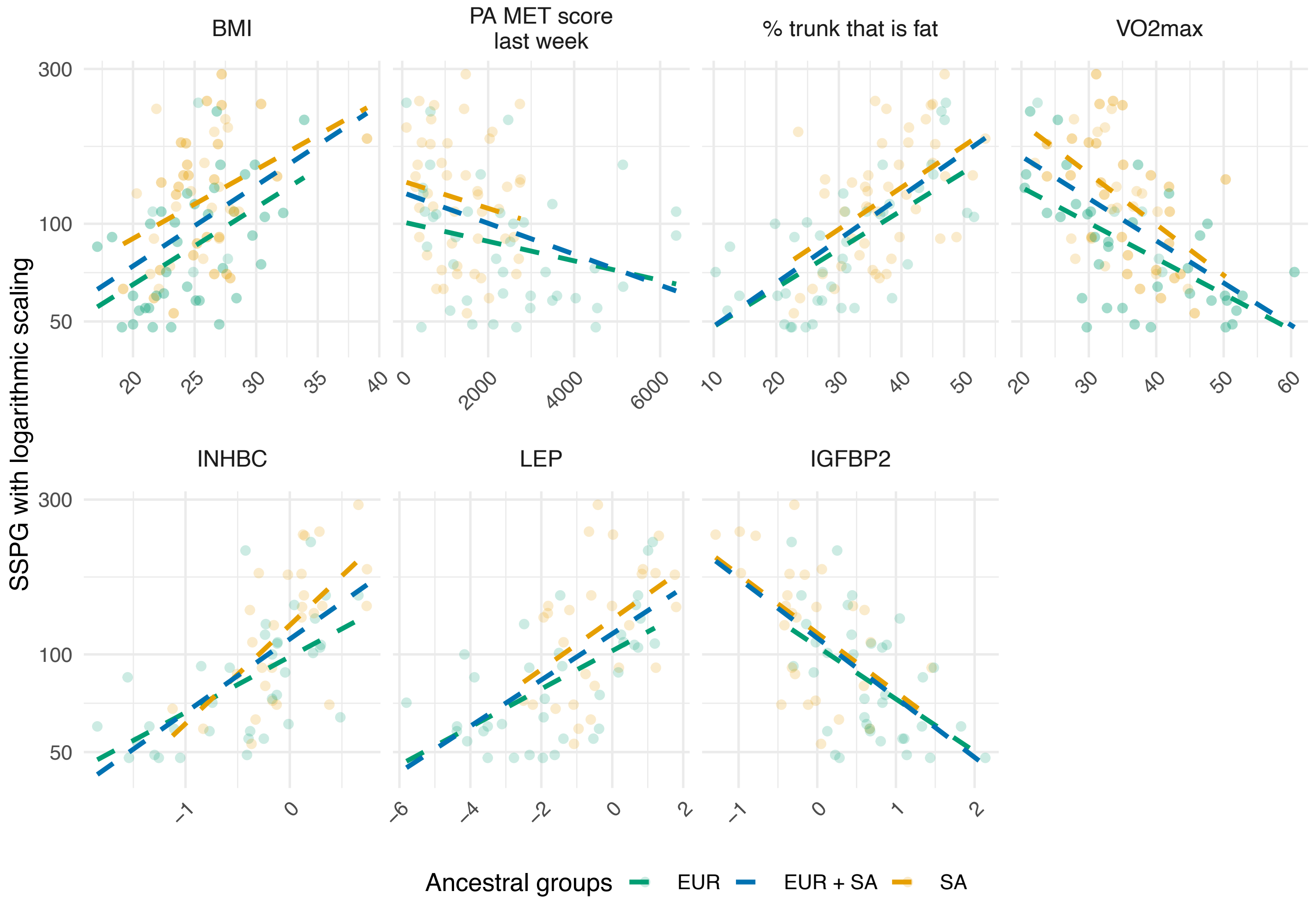


**Supplementary Figure 2. Scatter plot between SSPG and candidate predictors for EUR and SA participants.** Data points were colored according to ancestral group (green for EUR, orange for SA), with y-axes on logarithmic scale. Dashed lines show linear regression fits, using the same colors as the data points, while the blue dashed lines show the linear regression fit for both ancestry groups combined, without any additional covariates.

**Tables**

**Supplementary Table 1. Proteins and their full names with UniProt IDs**

| UniProt ID | Protein | Full Name |
|---|---|---|
| Q9Y653 | ADGRG1 | Adhesion G Protein-Coupled Receptor G1 |
| P12104 | FABP2 | Fatty Acid Binding Protein 2 |
| P15090 | FABP4 | Fatty Acid Binding Protein 4 |
| P08833 | IGFBP1 | Insulin Like Growth Factor Binding Protein 1 |
| P18065 | IGFBP2 | Insulin Like Growth Factor Binding Protein 2 |
| P55103 | INHBC | Inhibin Subunit Beta C |
| P06756 | ITGAV | Integrin Subunit Alpha V |
| P41159 | LEP | Leptin |
| P06858 | LPL | Lipoprotein Lipase |
| Q9BZR6 | RTN4R | Reticulon 4 Receptor |
| Q96PL1 | SCGB3A2 | Secretoglobin Family 3A Member 2 |

**Supplementary Table 2. Coefficient of ancestry and adjusted R2 values from regression analysis of log(SSPG) with selected groups of variables, adjusted for age, sex and ancestral group**

| Model | SAs vs EURs | | | Adjusted $R^2$ | | | Residual SE | | |
|---|---|---|---|---|---|---|---|---|---|
| | Beta | SE | P | Combining ancestral groups | SA only | EUR only | Combining ancestral groups | SA only | EUR only |
| Base model (age, sex) | 0.471 | 0.125 | 4.10E-04 | -0.026 | -0.045 | 0.069 | 0.489 | 0.486 | 0.414 |
| Conventional measures of adiposity and physical activity (BMI and PA MET scores last week) | 0.286 | 0.119 | 0.019 | 0.320 | 0.060 | 0.407 | 0.398 | 0.461 | 0.330 |
| Gold standard measure of adiposity (% trunk that is fat) and cardiorespiratory fitness ($VO_2$ max) | 0.142 | 0.116 | 0.228 | 0.452 | 0.357 | 0.368 | 0.357 | 0.381 | 0.341 |
| Plasma proteins (INHBC, LEP, IGFBP2) | 0.069 | 0.114 | 0.547 | 0.546 | 0.573 | 0.489 | 0.325 | 0.311 | 0.307 |

For model with sex, age and ancestral group, adjusted $R^2$=0.154 and residual SE=0.444. SAs, South Asians; EURs, Europeans; $R^2$, variance; SE, standard error; BMI, body mass index, PA, physical activity, MET: metabolic equivalent, VO2 max, amount of oxygen utilized during a cardiopulmonary exercise test with maximal effort, INHBC, Inhibin Subunit Beta C; LEP, Leptin; IGFBP2, Insulin Like Growth Factor Binding Protein 2
